# A high-fiber, low-fat diet improves the symptoms and metabolic profile of patients with Crohn’s disease

**DOI:** 10.1101/2024.08.30.24312853

**Authors:** Maria T. Abreu, Maria A. Quintero, Luis Garces, Hajar Hazime, Rose Killian, Katerina M. Faust, Payton Mendygral, Judith Pignac-Kobinger, Cristiana Mangarelli, Ana M. Santander, Irina Fernández, Norma Solis, Mailenys Ortega, Oriana M. Damas, Siobhan Proksell, David H. Kerman, Amar R. Deshpande, Jennifer Seminerio, Jana A.L. Hashash, Philip Harvey, Ingrid Barrera, Tracy Crane

## Abstract

**Background:** Crohn’s disease (CD) is characterized by intestinal inflammation. Diet is a risk factor for inflammatory bowel diseases such as CD and represents a promising adjunctive treatment, yet there are few well-controlled dietary intervention studies in CD patients. Fiber may have beneficial effects; however, most CD patients are told to avoid high-fiber foods. We conducted a longitudinal patient-preference study to examine the effect of a catered low-fat, high-fiber diet (the *Mi*-IBD diet) on CD symptoms, inflammation, and the microbiome.

**Methods:** CD patients (n=73) received one-time diet counseling (Group 1, n=23), the *Mi*-IBD diet (Group 2, n=26), or the *Mi*-IBD diet along with a healthy household control (Group 3, n=24 patient- HHC dyads). The *Mi*-IBD diet was catered for 8 weeks, and CD symptoms were recorded. Serum samples were collected to measure inflammatory marker levels and evaluate systemic changes via proteomic analyses (SomaScan Discovery v4.1 assays). Stool samples were collected to perform metabolomic analyses.

**Results:** At baseline, CD patients had a low-fiber, high-fat diet. One-time diet counseling did not result in dietary changes. In contrast, catering led to marked dietary changes in CD patients (increased fiber intake, decreased fat intake; all ps < 0.001) and high adherence rates (96%). Group 3 exhibited improvements in CD symptoms (PRO2 and sCDAI scores). Proteomic analysis revealed higher baseline serum levels of proinflammatory proteins (SAA and CRP) in CD patients than in HHCs; these levels decreased with the catered diet. The diet also improved fecal metabolites related to protein and energy metabolism as well as markers of oxidative stress and inflammation in CD patients.

**Conclusion:** A nonpharmacological approach involving a high-fiber, low-fat diet to manage CD was well tolerated, even by patients with fibrostenotic CD. These findings fill a gap in development of dietary recommendations for CD patients.

**Clinical trial registration:** This trial was registered at ClinicalTrials.gov (ID: NCT04213729).

## INTRODUCTION

Crohn’s disease (CD) is an inflammatory bowel disease (IBD) that affects the small intestine and colon. Advances in pharmacological therapy have occurred, but medical treatment does not restore normal intestinal function or result in endoscopic, histologic, or transmural healing in many patients^1^. Currently, non-pharmacological, evidence-based approaches for disease management (such as dietary interventions) are lacking. Although many epidemiological studies have linked diet to the development of CD, the certainty of evidence regarding the beneficial influence of specific diets in IBD is low^2^.

Several diets have been developed as non-pharmacological treatments for IBD, including Exclusive Enteral Nutrition (EEN), the Crohn’s Disease Exclusion Diet (CDED), the Mediterranean Diet (MD), the Specific Carbohydrate Diet (SCD), and the low-FODMAP diet^3,4^. Notably, among these diets, only CDED involves minimizing the consumption of animal fat; in the other diets, fat content is not regulated (e.g., SCD)^5^ or highly variable (e.g., EEN formulas can vary in fat content from 0% to 52.5%)^6^. These diets have been demonstrated to alleviate IBD symptoms and reduce inflammatory markers^3,4^. However, as mentioned above, the certainty of evidence is poor; thus, further well-controlled intervention studies are needed. Additionally, the adherence of CD patients to these diets is only moderate overall. Dietary adherence rates range from as low as 30% (EEN^7^) as high as 88% (low-FODMAP; at 4 weeks^8^). Thus, developing techniques to improve dietary adherence in CD patients is key for achieving better outcomes. One such approach is catering and delivering meals directly to participants; this technique was found to increase rates of adherence to the SCD or the MD by approximately 20%^9^. Additionally, family- or friend-based dietary interventions have reported greater weight loss among overweight individuals with greater dyadic support^10^. Thus, providing catered food for the entire household may improve CD patient adherence rates.

In a previous diet study of ulcerative colitis (UC) patients with quiescent disease, we discovered that patients typically consumed a diet that was high in fat and very low in fruits and vegetables^11^. Little is known about the effects of diet interventions focusing on fat and fiber intake on the management of CD. High fat intake may increase risk of disease activity in CD patients^12^, and saturated fatty acids (SFAs) act as non-microbial Toll-like receptor 4 (TLR4) agonists, triggering inflammation^13^. In general, increased fiber intake is associated with salutary effects, in part through its effects on the microbiome; fiber leads to the generation of short-chain fatty acids (SCFAs), which may have anti-inflammatory effects^14^. However, most patients with CD are told to eat a low-fiber diet due to concern that fiber may cause intestinal obstruction or worsen symptoms of diarrhea, especially in patients with a history of fibrostenotic disease^15^.

Diet also influences the functional microbiome, altering the bacterial metabolites produced in the gut. IBD patients consistently exhibit altered fecal metabolomic profiles from those of healthy controls, with depleted fatty acid levels^16^. In healthy individuals, changes in diet (consumption of an isocaloric MD) were demonstrated to alter the metabolome, enhancing microbial carbohydrate metabolism and reducing bile acid production^17^. Thus, it is reasonable to expect that a diet intervention may improve microbiome function in IBD patients, potentially rescuing some of the alterations observed in IBD.

In the present study, our primary hypothesis was that CD patients adhering to a high-fiber, low-fat diet (the *Mi*-IBD diet) would exhibit decreased symptoms and inflammation compared to those receiving diet counseling only. Our secondary hypothesis was that catering the dietary intervention would improve both dietary adherence (i.e., decreased fat intake and increased fiber intake) and our ability to rigorously examine the microbiome and metabolome, increasing experimental control. We further predicted that the Mi-IBD diet would improve microbiome function, shifting the CD patient metabolomic profile toward that of healthy individuals. The study design was novel in that we used a patient-preference approach and included only CD patients living with an adult with whom they shared meals. Patients chose to receive one-time diet counseling or catered meals (20% of total calories from fat and 14 g of fiber/1,000 calories per day) for eight weeks. Our results demonstrated that a low-fat, high-fiber diet is well tolerated in CD patients, leading to improvements in CD symptoms. The inclusion of a household control group gave us further power to understand how a reasonably simple diet affected microbiome function as measured by metabolomics as well as systemic proteomic markers. Our results demonstrate improvements in the CD patients in systemic inflammatory markers and improved fecal metabolites related to protein and energy metabolism, oxidative stress, and inflammation.

## MATERIALS & METHODS

### Study design

This patient-preference study had a prospective, longitudinal design (**Supplementary** Figure 1). Participants were recruited by physicians during routine visits or by telemedicine through the University of Miami. Participants from other states were also recruited through clinicaltrials.gov (ID: NCT04213729). Recruitment and study completion occurred between January 7, 2020, and July 10, 2023 (see the Supplementary Materials for further details). The study was approved by the University of Miami Institutional Review Board (protocol 20190548). All study participants provided written informed consent either online or on a paper form before participating in the study. All authors had access to the study data and reviewed and approved the final manuscript.

### Patient enrollment and group allocation

Patients diagnosed with mild to moderate CD were eligible for inclusion if they lived with another adult with whom they generally shared meals (see the Supplementary Materials for all inclusion and exclusion criteria). Patients chose one of three groups: Group 1 (n=23) received a one-time diet counseling session, Group 2 (n=26) received catered food (the *Mi*-IBD diet) for an eight-week period, and Group 3 received catered food along with healthy household controls (HHCs) who were free of gastrointestinal disease (n=23 dyads). These HHCs were included as an important control for the effect of the same diet (the Mi-IBD diet) on a healthy individual compared to a CD patient while maintaining a similar environment (the shared household).

### Demographic and clinical characteristics

Information on age, gender, race, ethnicity, birth country, marital status, and anthropometric measurements was collected from participants at baseline. Additional clinical characteristics were assessed in CD patients: disease location, Montreal classification, current medication use, previous medication use, and history of gastrointestinal surgery.

### Characteristics of the *Mi*-IBD diet

The *Mi*-IBD diet consisted of 20% of total calories from fat, an omega-6/omega-3 fatty acid ratio of approximately 1:1, and 14 g of fiber per 1,000 calories consumed. The catered meals included breakfast, lunch, dinner, and snacks and were catered by national catering companies that used organic, minimally processed, high-quality ingredients (see the Supplementary Materials).

### Assessments of diet intake and CD symptoms

Participants completed the Automated Self-Administered 24-hour Dietary Assessment Tool (ASA24)^18^ at baseline and week 8 (Supplementary Figure 1). This self-report tool is used to assess micronutrients and macronutrients consumed based on participant recall of foods consumed over the past 24 hours. From the ASA24 data, the healthy eating index (HEI) scores were calculated. The HEI is used to measure diet quality relative to the United States Department of Agriculture (USDA) recommendations in the *Dietary Guidelines for Americans*^19^ and evaluates 13 dietary components, including total fruits, total vegetables, whole grains, refined grains, and fatty acids. Higher scores reflect higher dietary quality.

Three validated instruments were administered each month to assess CD disease activity: the Patient-Reported Outcomes-2 (PRO2), Short Crohn’s Disease Activity Index (sCDAI), and Harvey-Bradshaw Index (HBI).

### Measurement of inflammatory markers

Serum samples and morning stool samples at baseline and week 8 were collected to measure C-reactive protein (CRP), serum amyloid A (SAA), and calprotectin. CRP levels were measured using a Human CRP/Reactive Protein ELISA PicoKine Kit (Catalog #EK1316; Boster Biological, Pleasanton, CA). SAA determination was performed using a Human Serum Amyloid A DuoSet ELISA Kit (Catalog #DY3019-05; R&D Systems, Minneapolis, MN). Fecal calprotectin was measured with a BUHLMANN fCAL ELISA Kit (Catalog #EK-CAL2; BUHLMANN Laboratories, Amherst, NH). Calprotectin was extracted with a CALEX Cap Device (Catalog #B- CALEX-C200; BUHLMANN Laboratories).

### Serum proteomics analysis to assess systemic markers

Proteomic analysis of serum samples was performed by SomaLogic (Boulder, CO) using the SomaScan Discovery v4.1 assay, which can detect 5,000 human protein analytes. Specifically, serum samples from 10 dyads (CD patient from Group 3 + HHC) and 10 CD patients from Group 1 were analyzed at baseline and week 8. These participants were randomly selected from among the participants with blood samples available for both baseline and week 8.

### Untargeted metabolomic analysis of stool samples

Stool samples were sent to the Targeted Metabolomics and Proteomics Laboratory at the University of Alabama at Birmingham for untargeted metabolomics via liquid chromatography– mass spectrometry (LC-MS)/MS global analysis. Data were normalized (total ion current) and subjected to Pareto scaling prior to univariate and multivariate statistical analysis, hierarchal heatmap clustering, and pathway analysis using MetaboAnalyst version 5 (http://www.metaboanalyst.ca).

### Outcome measures

The outcome measures were improvements in clinical symptoms in patients consuming an Mi-Diet. The secondary outcome measure was a change in stool metabolomic profile of CD patients towards that of healthy subjects.

### Statistical analysis

Group differences were detected using ANOVAs. Bartlett’s test of homogeneity of variance was performed to detect differences in variance among groups; if the standard ANOVA assumption of equal variance was violated, Welch’s ANOVA was utilized. Post hoc t-tests with Bonferroni correction were conducted if a significant difference was identified. Additionally, paired t-tests were used to detect changes from baseline within groups. Analyses were performed and figures were generated with GraphPad Prism version 10.1.2 (GraphPad Software, Boston, MA). The threshold for statistical significance was set at p ≤ 0.05. HEI components and dietary macronutrients other than fat and fiber intake are reported without adjustment for multiple comparisons in the **Supplementary Materials**.

## RESULTS

### Demographic and clinical characteristics of participants

Overall, 115 participants were enrolled in the study, and 73 CD patients and 24 HHCs completed all study requirements (see **Figure 1**). The CD patients consisted of equal numbers of men (47.9%) and women (52.1%). The average age of CD patients was 36.8 ± 12.4 years, and the average disease duration was 11.1 ± 9.9 years. Approximately half of the patients (55.2%) self-identified as Hispanic/Latino. Most CD patients were in clinical remission according to their mean HBI score (HBI score: 3 ± 2.9)^20^. There were no significant differences in HBI, PRO2, or sCDAI scores among the groups at baseline. The levels of inflammatory markers (fecal calprotectin, CRP, and SAA) were low, consistent with quiescent CD. One third of patients (32.4%) had fibrostenotic disease; patients with obstructive symptoms were not enrolled (see **Supplementary** Figure 2). Further details regarding the included participants are shown in **Table 1**.

**Figure 1.**
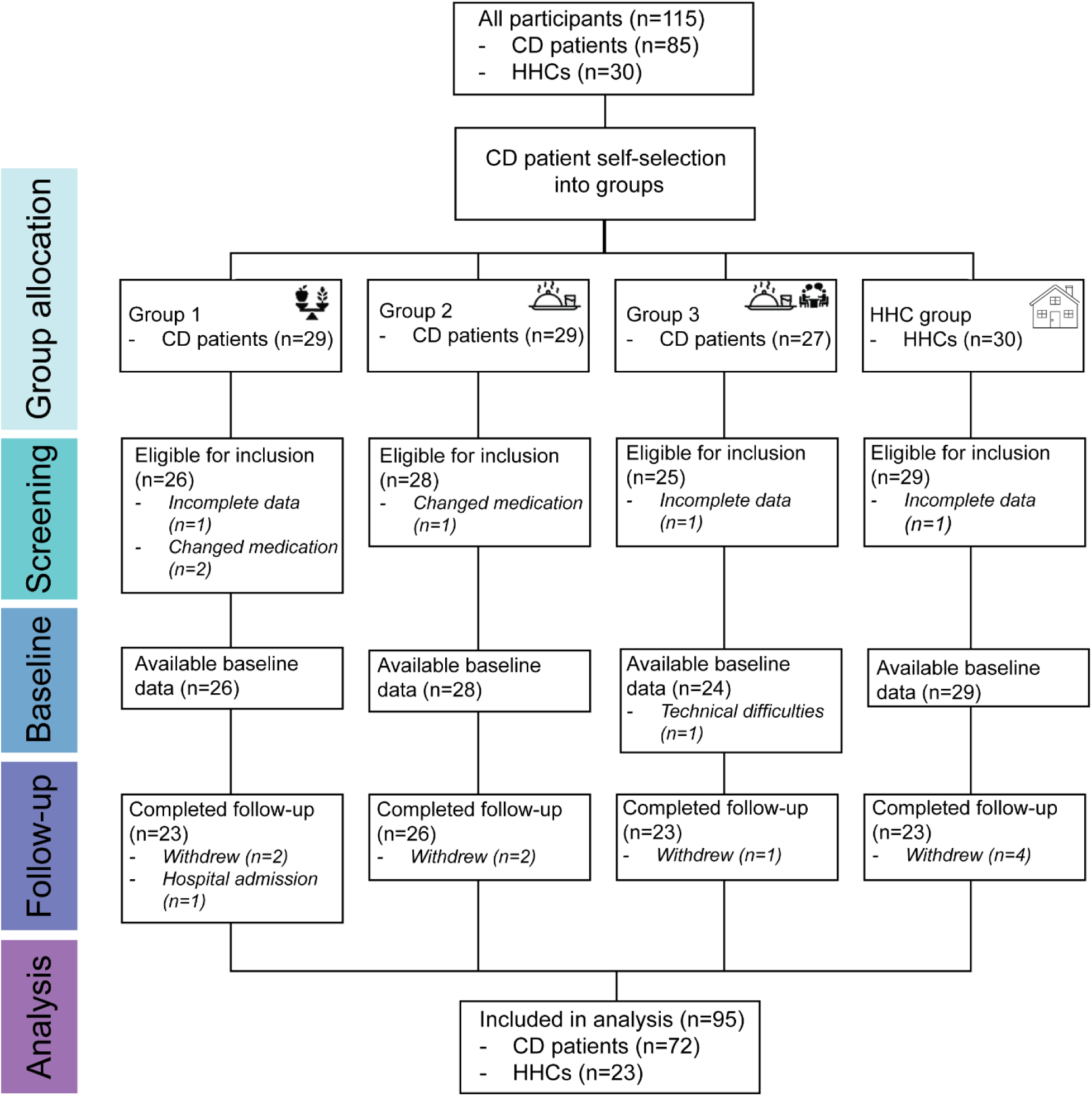
Participant enrollment and group allocation flowchart. Abbreviations: CD = Crohn’s disease, HHC = healthy household control.

**Table 1.** Demographic and clinical characteristics of the included participants (n=95).

|  | Group 1<br>(n=23) | Group 2<br>(n=26) | Group 3<br>(n=23) | HHCs<br>(n=23) |
| --- | --- | --- | --- | --- |
| Age, median (range) | 37 (20-59) | 37 (21-66) | 37 (23-63) | 36 (21-63) |
| Female, n (%) | 14 (60%) | 12 (46%) | 11 (45.8%) | 14 (58.3%) |
| Race, n (%) |  |  |  |  |
| White | 20 (86.9%) | 24 (92%) | 22 (91%) | 19 (79.2%) |
| Black | 1 (4.3%) | 0 | 0 | 0 |
| Asian | 0 | 0 | 0 | 1 (4.2%) |
| Other | 0 | 0 | 0 | 3 (12.5%) |
| Unknown | 2 (8.6%) | 2 (7.6%) | 2 (8%) | 1 (4.2%) |
| Ethnicity, n (%) |  |  |  |  |
| Hispanic | 9 (39%) | 12 (46%) | 14 (58%) | 18 (75%) |
| Non-Hispanic | 14 (60%) | 14 (53.8%) | 9 (37%) | 6 (25%) |
| Unknown | 0 | 0 | 1 (4%) | 0 |
| Birth country, n (%) |  |  |  |  |
| USA | 15 (65%) | 11 (42%) | 15 (62%) | 10 (41.7%) |
| Other | 8 (23%) | 15 (57%) | 9 (37%) | 14 (58.3%) |
| Marital status, n (%) |  |  |  |  |
| Never married | 6 (26%) | 8 (30%) | 6 (25%) | 3 (12.5%) |
| In a relationship | 4 (17%) | 2 (7.6%) | 2 (8.3%) | 3 (12.5%) |
| Married | 11 (47.8%) | 14 (53.8%) | 16 (66%) | 18 (75%) |
| Divorced/separated | 1 (4.4%) | 2 (7.6%) | 0 | 0 |
| Baseline BMI (kg/m<sup>2</sup>), mean (SD) | 26.8 ± 6 | 26.5 ± 5.9 | 27.08 ± 6.18 | 28.6 ± 5.6 |
| Age at diagnosis, median (range) | 25 (9-45) | 24.9 (10-58) | 24.51 (12-58) | -- |
| History of GI surgery, n (%) | 8 (34%) | 9 (34.6%) | 11 (45.8%) | -- |
| HBI score, mean ± SD | 2.75 ± 2.2 | 3.1 ± 3.2 | 3.25 ± 3.1 | -- |
| Disease location |  |  |  |  |
| L1 – terminal ileum | 7 (30%) | 6 (23%) | 11 (45%) | -- |
| L2 – colon | 2 (8.6%) | 2 (7.6%) | 2 (8.3%) | -- |
| L3 – ileocolon | 14 (60%) | 17 (66%) | 10 (41%) | -- |
| L4 – upper GI | 0 | 1 (3.8%) | 1 (4.1%) | -- |
| Montreal classification |  |  |  |  |
| B1 – inflammatory | 15 (65%) | 10 (38%) | 13 (54%) | -- |
| B2 – fibrostenotic | 7 (30%) | 12 (46%) | 6 (25%) | -- |
| B3 – perforating | 1 (4.3%) | 4 (15%) | 5 (20%) | -- |
| Current medication use*, n |  |  |  | -- |
| Aminosalicylates | 1 | 1 | 0 | -- |
| Steroids | 0 | 0 | 0 | -- |
| Immunomodulators | 1 | 3 | 4 | -- |
| Biologics | 18 | 23 | 18 | -- |

| Previous medication use*, n |  |  |  |  |
| --- | --- | --- | --- | --- |
| Aminosalicylates | 11 | 10 | 2 | -- |
| Steroids | 11 | 4 | 13 | -- |
| Immunomodulators | 5 | 8 | 7 | -- |
| Biologics |  |  |  | -- |
| 1 biologic | 3 | 6 | 5 | -- |
| 2 biologics | 1 | 2 | 1 | -- |
| 3 biologics | 0 | 2 | 1 | -- |
| >3 biologics | 0 | 1 | 0 | -- |

\*Percentage values are not provided because the categories are not mutually exclusive.
*Note.* Abbreviations: BMI = body mass index, CD = Crohn's disease, HBI = Harvey-Bradshaw Index, GI = gastrointestinal, SD = standard deviation.

### The baseline diet of CD patients is low in fiber and high in fat

The United States Department of Agriculture (USDA) recommends consuming a diet with 14 g of fiber per 1,000 calories per day^21^ and less than 10% of calories from saturated fat^22^. Baseline dietary consumption was assessed with the ASA24, and these data were used to derive HEI scores. At baseline, the ASA24 results indicated that CD patients consumed approximately 10 g of fiber per 1,000 calories, 36.8% of calories from fat, and >10% of calories from saturated fat. All three CD patient groups had similar baseline levels of macronutrient intake (protein, total fat, saturated fat, dietary cholesterol, fiber, sugar, and the ratio of omega 6 to omega 3 fatty acids) and similar HEI scores (**Supplementary Table 1**). The average HEI total score of the general US population is 55.3^23^; our CD patients had average values ranging from 49.8 to 53.3. By definition, the ideal HEI total score is 100, reflecting complete alignment with dietary recommendations. These data demonstrate that CD patients with quiescent disease consume a diet that is not consistent with the current USDA nutritional guidelines^24^.

### Catering meals achieves the nutritional goals of the Mi-Diet in CD patients and household controls whereas one-time diet counseling does not

We assessed dietary adherence during the study period (eight weeks) by examining data logged in a daily food diary (Nutrihand). All groups provided with catered meals (Group 2, Group 3, and the HHC group) consumed approximately 80% of the provided food. Almost all patients (96%) also reported that they *always* or *usually* ate the catered food (see **Supplementary Materials** for further details). Further details regarding the intake of specific nutrients in each group at baseline and week 8 are shown in **Supplementary** Figure 3.

We further examined macronutrient intake according to the ASA24 to determine whether participants receiving the catered *Mi*-IBD diet achieved the dietary intake targets (**Figure 2**). One-time diet counseling did not result in dietary changes. In contrast, catering led to significant decreases in total fat intake and the percentage of calories derived from saturated fat as well as significant increases in fiber intake and protein intake (all ps < 0.001). Thus, our intervention achieved our primary aim of improving dietary adherence, leading to reduced fat intake and increased fiber intake.

**Figure 2.**
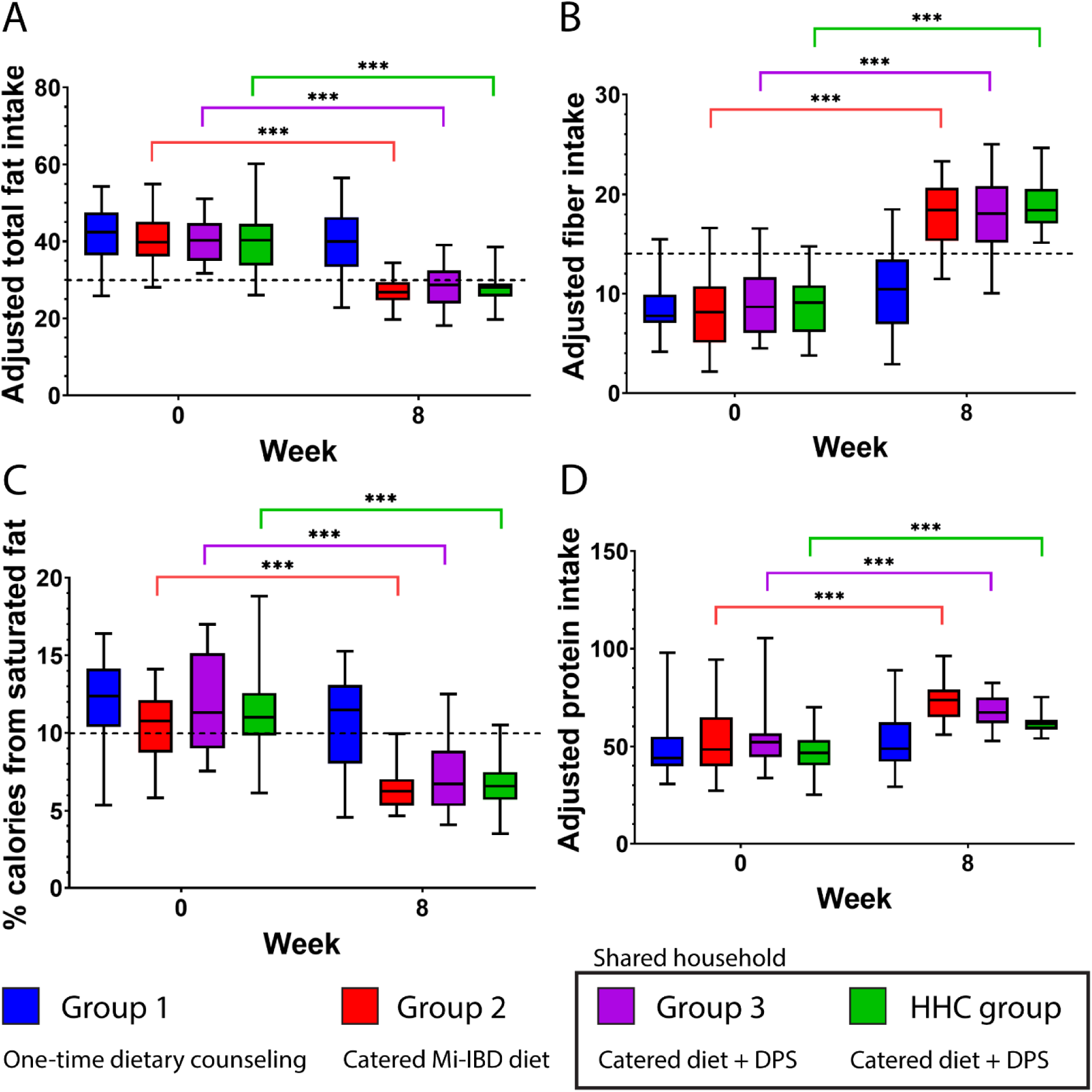
The *Mi*-IBD diet decreased fat intake and increased fiber and protein intake. A) Fat intake, B) fiber intake, C) percentage of calories from saturated fat, and D) protein intake in all groups. Dashed lines indicate recommended dietary intake amounts. All intakes are adjusted for total daily intake (grams per 1,000 calories consumed), except for C. Data shown are from the ASA24. Abbreviations: HHC, healthy household control. ASA24 data were analyzed with paired t-tests with Bonferroni correction. * p < 0.05, ** p < 0.01, *** p < 0.001 (asterisks reflect adjusted p values).

Participants who received the catered food met the target threshold of consuming 14 g of fiber per 1,000 calories at week 8, while those who did not receive the catered food did not meet this threshold. Although the percentage of calories derived from fat at week 8 in the groups that received the catered food remained above the target (<20% of calories from fat), these values were substantially improved from the baseline values and from those of Group 1. The percentages of calories from saturated fat significantly decreased in the groups that received the catered meals, matching the USDA nutritional guidelines threshold (of 10% or less^22^); no such decrease was observed in Group 1.

Finally, we examined dietary quality among the groups in terms of changes in HEI scores from baseline to week 8 (**Supplementary** Figure 5). Group 1 did not exhibit any improvements in HEI scores over time. In contrast, overall dietary quality (HEI total scores) improved from baseline to week 8 in Group 2, Group 3, and the HHC group. Specifically, the catered meals led to significantly increased consumption of total vegetables, greens and beans, total fruit, whole fruit, and whole grains and significantly decreased consumption of refined grains and saturated fat.

### The *Mi*-IBD diet improved CD symptoms

We next investigated whether consumption of the *Mi*-IBD diet led to improvements in the symptoms of CD patients. Specifically, we compared HBI, PRO2, and sCDAI scores between baseline and week 8 (**Figure 3**; **Supplementary Table 2**). Groups 1 and 2 did not exhibit any significant changes in HBI, PRO2, or sCDAI scores. In contrast, Group 3 exhibited improvements in both PRO2 and sCDAI scores from baseline to week 8. In other words, receiving the catered food along with a family member led to improvements in CD symptoms.

**Figure 3.**
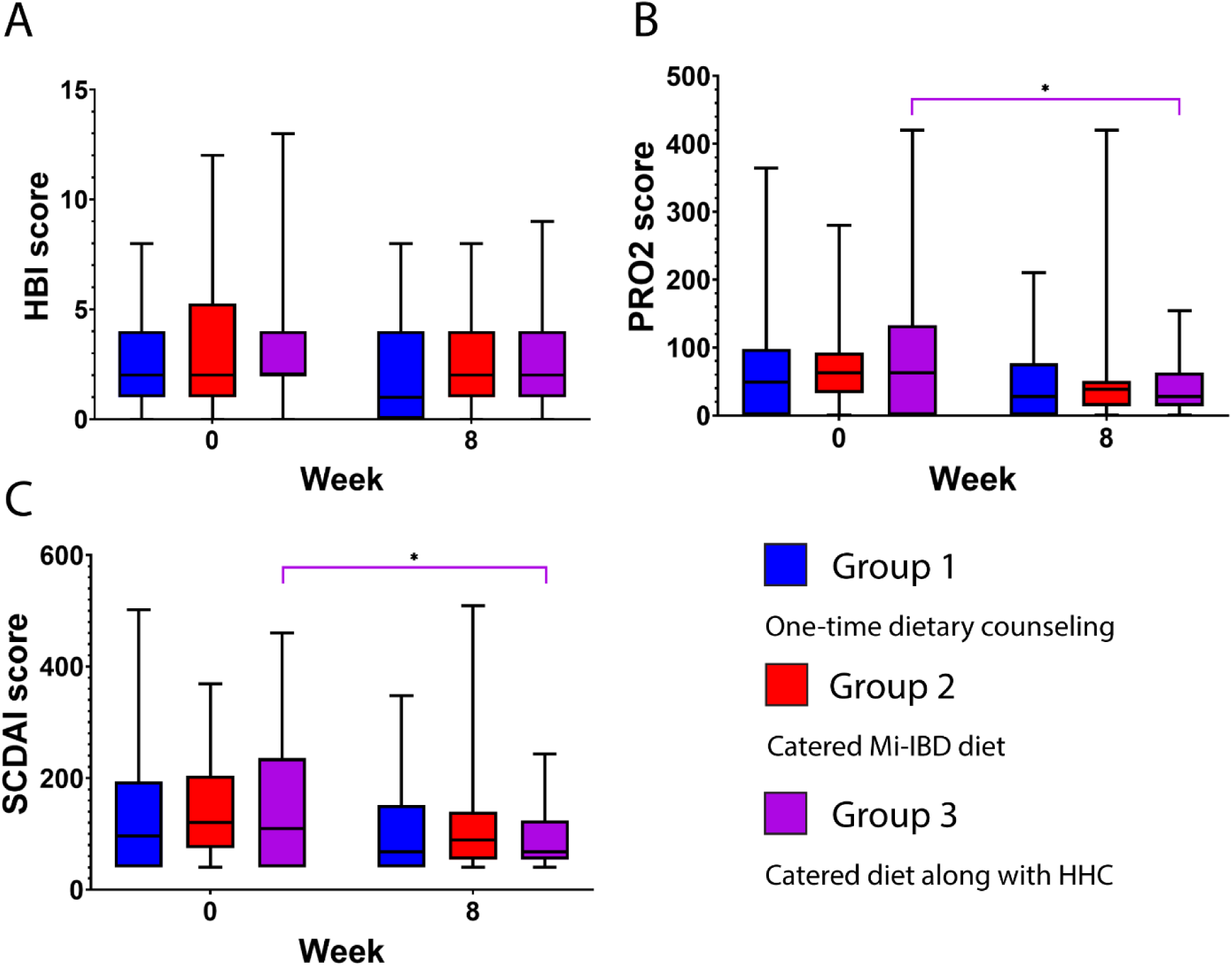
Receiving the *Mi*-IBD diet along with a family member led to improvements in CD symptom severity. A) HBI values, B) PRO2 scores, and C) sCDAI scores in the CD patient groups. Abbreviations: CD = Crohn’s disease, HBI = Harvey-Bradshaw index, HHC, healthy household control, PRO2 = Patient-Reported Outcomes-2, sCDAI = Short Crohn’s Disease Activity Index. Data were analyzed with paired t-tests with Bonferroni correction. * p < 0.05 (asterisks reflect adjusted p values).

Importantly, consumption of the *Mi*-IBD diet did not lead to an increase in abdominal pain (PRO2 scores) or CD symptom severity (HBI scores). We also examined the subset of CD patients with fibrostenotic disease; these patients are typically advised to avoid fiber intake. Patients with a fibrostenotic phenotype present in all groups exhibited no changes in HBI, PRO2, or sCDAI scores from baseline to week 8 (all ps > 0.11). Thus, the *Mi*-IBD diet, which is high in fiber, did not lead to increased CD symptoms among patients with fibrostenotic disease.

### The *Mi*-IBD diet differentially impacted serum protein levels and pathway activity in CD patients and HHCs

CD patients enrolled in this study generally had mild symptoms or were in symptomatic remission with low levels of inflammatory markers at baseline. Inflammatory marker levels in stool and serum samples were assessed at baseline and week 8 (**Supplementary Table 3**). CRP levels did not exhibit significant changes from baseline to week 8 in any group (all ps > 0.24). Similarly, fecal calprotectin levels did not exhibit significant changes from baseline to week 8 in any group (all ps > 0.15); however, there was a nonsignificant trend toward a difference in Group 2 (t(24) = -1.87, p = 0.07). SAA levels did not significantly change from baseline to week 8 in any group (all ps > 0.1). Overall, these data suggest that the *Mi*-IBD diet did not worsen inflammation.

After assessing established inflammatory markers, we wanted to have an agnostic view of the differences in systemic effects of the diet in patients with CD versus household controls.

We thus focused on the Group 3 CD patient-HHC dyads, allowing us to compare changes in CD patients with those in matched HHCs. For these analyses, we performed SomaLogic proteomics in ten randomly selected dyads.

At baseline, CD patients had higher levels of pro-inflammatory proteins (SAA1 and CRP), immunomodulatory proteins (CLEC12A) and IgA) than HHCs (**Figure 4A**). Consumption of the Mi-IBD diet reduced levels of leptin (a protein derived from adipocytes^25^) and C1QBP (a protein that impairs goblet cell formation in IBD patients^26^ and plays a proinflammatory role^27^) in CD patients (**Figure 4B)**. In contrast, the *Mi*-IBD diet increased levels of proteins important for maintenance of the intestinal epithelium (RAB21 and RAB74)^28^.

**Figure 4.**
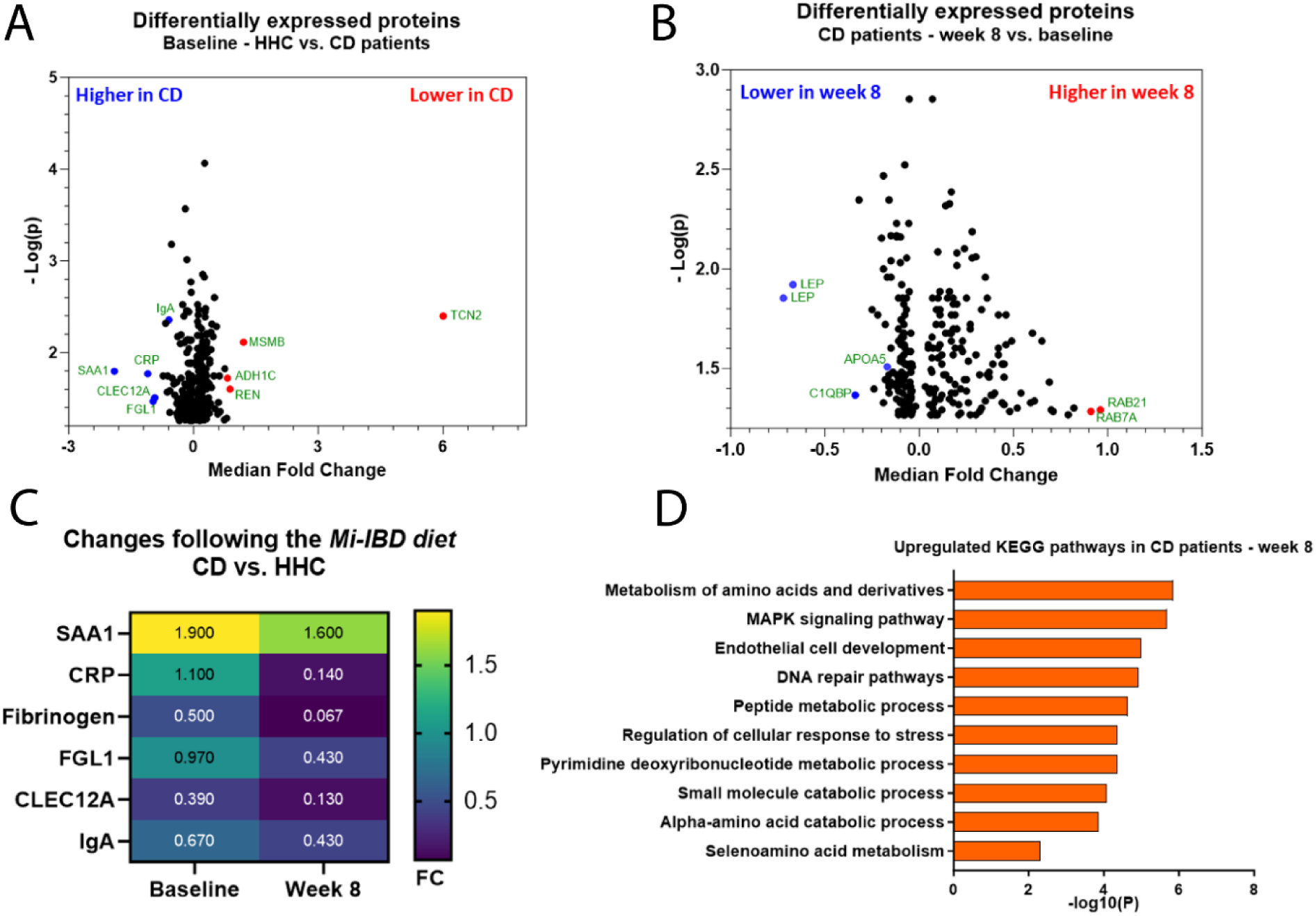
Proteomic analysis of serum samples from CD patient-HHC dyads (n=10). A) Volcano plot showing differentially expressed proteins at baseline in the sera of HHCs and CD patients. B) Volcano plot showing differentially expressed proteins in the sera of CD patients at week 8 compared to baseline. C) Heat map showing the change in select differentially expressed proteins between CD patients and HHCs at baseline and week 8. D) KEGG pathway analysis of differentially expressed proteins in CD patients at week 8 compared with baseline. CD patient composition: 60% ileal only, 40% colonic involvement. Abbreviations: CD = Crohn’s disease, HHC = healthy household control.

We compared protein levels in HHCs and CD patients before and after the diet. Consumption of the *Mi*-IBD diet by CD patients reduced levels of inflammatory and immunomodulatory markers, approaching those of HHCs (**Figure 4C**). However, these diet-induced changes were unable to completely eliminate the differences between CD patients and HHCs; inflammatory and immunomodulatory markers remained higher in CD patients.

Finally, we conducted pathway analysis within CD patients. Consumption of the *Mi*-IBD diet significantly upregulated protein expression in pathways associated with amino acid metabolism, fatty acid metabolism, endothelial cell development, and cellular responses to DNA damage (**Figure 4D**). Together, these changes in serum protein levels may reflect systemic diet-induced improvements in inflammation and metabolism in CD patients.

### Consuming the Mi-IBD diet changed the fecal metabolome of CD patients

To gain insight into the impact of diet modification on microbiome function, we analyzed the fecal metabolome of our patients before (week 0) and after (week 8) consuming the catered diet. Metabolomic analysis at week 8 revealed a marked separation in the metabolome of groups that received the catered diet (Group 2, Group 3, and the HHC group) compared to the group that did not receive catered food (Group 1) (**Figure 5A**). Examination of specific fecal metabolites revealed that CD patients who received the catered diet (Groups 2 and 3) show significant metabolomic improvements compared to patients who did not receive the diet (Group 1), underscoring the Mi-IBD diet’s role in enhancing metabolomic health (**Figure 5B**). The Mi-IBD diet improved fecal metabolites related to protein and energy metabolism as well as markers of oxidative stress and inflammation in Groups 2 and 3; these markers remained unchanged in Group 1. Notably, although both Group 2 and Group 3 received the catered diet and showed improvements, Group 3 displayed more pronounced changes, possibly due to the shared household environment with HHCs consuming the same diet.

**Figure 5.**
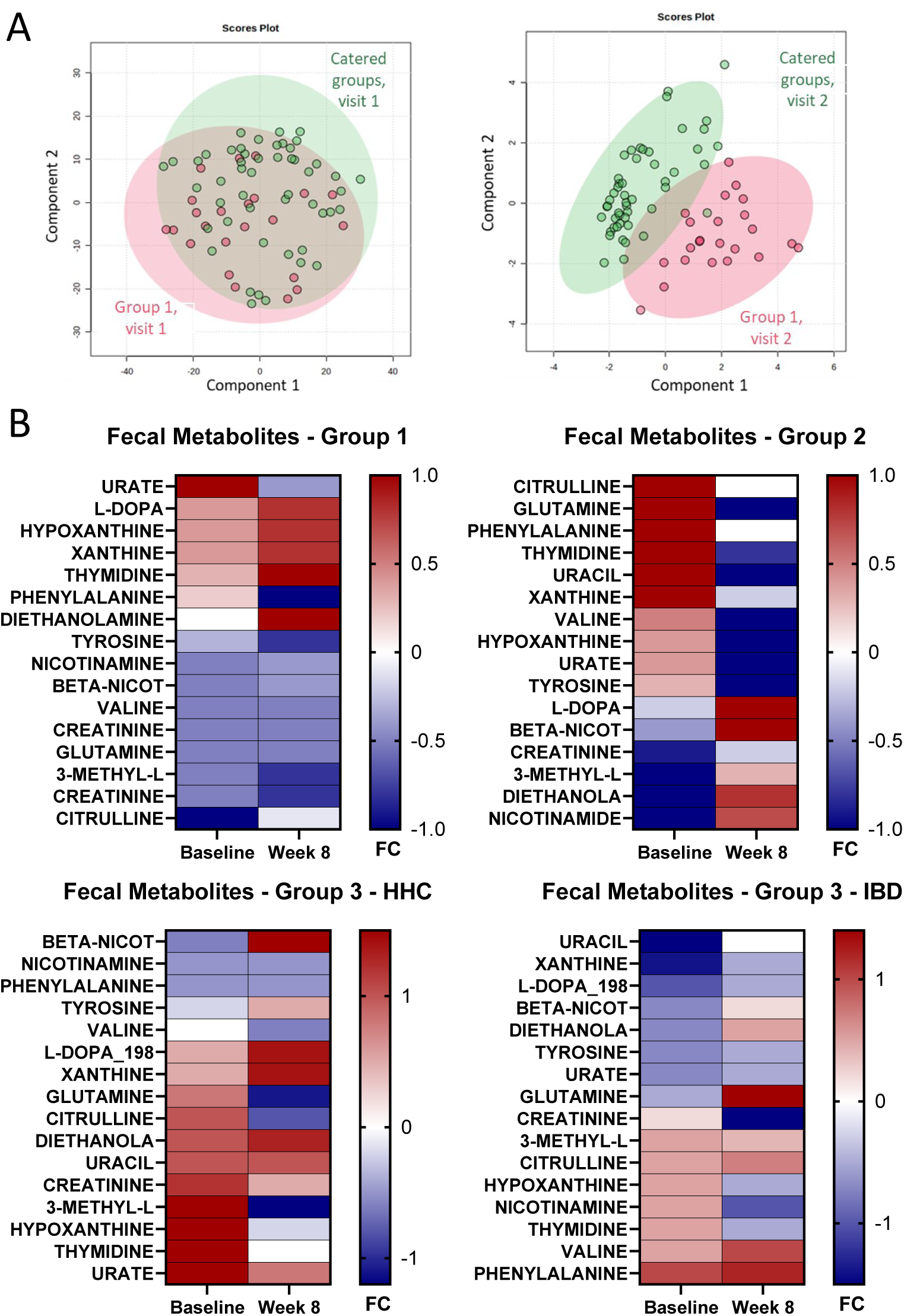
Metabolomic analysis of stool samples from CD patient-HHC dyads. A) Principal component analysis (PCA) of untargeted fecal metabolites from CD patients (Group 1: n=23 vs. Groups 2 & 3: n=49). B) Heat map showing differentially clustered metabolites from the stool of HHCs and CD patients before and after the LFD intervention (Group 1: n=23, Group 2, n=26, Group 3: n=23, HHCs: n=23). Abbreviations: CD = Crohn’s disease, HHC = healthy household control, FC = fold change.

Next, we leveraged our data to examine whether the impact of the Mi-IBD diet varied between CD patients and HHCs living in the same household. We found that HHCs showed more stable metabolomic profiles than CD patients (Group 3), who exhibited more variable responses. This highlights the diet’s differential impacts in healthy vs. disease states. Both groups showed improvements in metabolites related to energy metabolism, but Group 3 exhibited more significant reductions in oxidative stress markers than the HHCs, suggesting that patients had a stronger reduction in inflammation in response to the dietary intervention.

## DISCUSSION

Patients with IBD often want to receive dietary counseling, with 80-89% of CD patients asking their physicians for dietary advice^29^. However, it remains unclear which diets are beneficial for IBD, leading to confusion and reduced food-related QoL in patients. As a result, many patients with IBD develop avoidant/restrictive food intake disorder (ARFID)-like conditions and have an overly restrictive diet^30^. At the same time, patients who receive dietary counseling are rarely completely adherent to this advice, with low levels of dietary adherence reported by previous studies. In this study, we attempted to answer two important questions: 1) Does catering a low-fat, high-fiber diet improve dietary adherence? 2) Does this diet improve CD symptoms, inflammation, and microbiome function?

To our knowledge, this is the first patient-preference study of diet to be completed in patients with CD. We believe that patient self-selection into groups is a necessary aspect of this study because the design involved the participation of household members (Group 3 CD patient- HHC dyads); thus, we could not simply randomly assign CD patients to groups without considering the willingness of household members to participate in this study. Importantly, the groups were well matched for baseline characteristics relevant to the study, such as baseline diet, disease activity, and QoL scores. Moreover, we view the patient-preference approach as a strength of our design, as it enabled us to achieve high adherence rates (and thereby excellent experimental control of diet). Another strength of our study was enrollment of a diverse patient population: almost half of our CD patients were Latino. Therefore, our results can be extrapolated to other cultures.

Our findings shed light on the importance of catering to achieve high adherence rates in dietary interventions. We included a group that received one-time diet counseling, a group that received catered food for themselves only, and a group that received catered food along with a household member. Notably, while patients who received the catered food had very high adherence rates (96%), catering food for a patient and their household member did not lead to further improvements in adherence (i.e., no differences in adherence between Groups 2 and 3), possibly because of a ceiling effect. This aligns with previous findings in healthy people that both individual and household-based interventions successfully reduced fat intake and that there were no significant differences in the reduction in fat intake between the two intervention types^31^. Overall, CD patients want to receive dietary advice, but we very clearly showed that they do not adhere to this information once provided. Unfortunately, one-time counseling sessions are the most common type of guidance provided by dieticians to CD patients. Dieticians who want to achieve sustained change should consider directing patients to catering companies that can provide food in line with physician recommendations.

In terms of the specific diet that we evaluated, we demonstrated that the *Mi*-IBD diet was well tolerated in patients with CD, including those with a fibrostenotic phenotype. Many patients with the fibrostenotic phenotype are counseled not to eat fiber^32^. Additionally, most CD patients at any given time are in remission^33^ but still avoid consuming fruits and vegetables^34^ due to their fiber content. However, when receiving a catered diet including high fiber intake, patients in Group 3 reported improvements in abdominal pain, bowel movements, and overall sCDAI scores. The sources of fiber in the *Mi*-IBD diet were whole grains, fruits, vegetables, legumes, and nuts and seeds. Patients reported enjoying the meals, particularly the ability to eat fruits and vegetables again; indeed, on the *Mi*-IBD diet, their fruit consumption nearly doubled.

We did not see overall biochemical improvements (e.g., in CRP or fecal calprotectin levels). This lack of change was likely due to the inclusion of patients who were on stable medications and doing well at the time of recruitment. Using the aptamer-based SomaLogic SomaScan Discovery v4.1 assay, we observed that CD patients, despite being in clinical remission, had elevated systemic markers of inflammation. These systemic inflammatory markers improved with the *Mi*-IBD diet intervention. We believe that our diet strategy (which we previously tested in UC patients^11^) can be added to the list of diets recommended for IBD patients, including EEN, CDED, MD, and SCD.

In addition to the direct impact of diet on CD symptoms, improved dietary quality is associated with broad health benefits, extending beyond IBD. First, there is growing recognition of shared features of metabolic syndrome and IBD^35^, with metabolic dysfunction-associated fatty liver disease (MAFLD) observed in 42% of IBD patients^36^. Second, higher visceral adiposity is associated with a decreased response to monoclonal antibody biologic therapies in IBD^37^. Thus, reduced fat intake (such as consuming the *Mi*-IBD diet) may reduce visceral adiposity and risk of MAFLD; reduced visceral adiposity may facilitate treatment response in IBD patients. Third, these patients also have higher risks of venous thromboembolism (VTE) and coronary artery disease^38^. IBD patients may be treated with Janus kinase (JAK) inhibitors (e.g., tofacitinib)^39^, but JAK inhibitors elevate the risk of cardiovascular complications such as VTE^40^. Thus, it is important to prescribe a healthy diet that minimizes cardiovascular risks in IBD patients. Our Mi-IBD diet improved fecal metabolites related to protein and energy metabolism as well as markers of oxidative stress and inflammation and can thus be recommended to CD patients.

### Limitations

This study has some limitations. First, we did not perform colonoscopies before and after the intervention to assess mucosal healing. Second, the use of self-selection (patient preference) for group allocation is a novel approach for this type of study. Although we examined potential group differences at baseline, we may not have captured additional factors, such as willingness of the dyadic partner to support the patient.

## Conclusion

A diet high in fiber and low in saturated fat is well tolerated and can be recommended to patients with CD. Catering this diet resulted in very high adherence rates, especially compared with the lack of dietary changes observed in response to one-time diet counseling. Our *Mi*-IBD diet, especially when combined with the participation of a household member, improved patient symptoms. We observed significant changes in the proteomic and metabolomic profile of CD patients, suggesting improvements in systemic inflammation, metabolism, and repair that, if sustained, could facilitate maintenance of remission and decrease the risk of metabolic syndrome. Our findings can be used to refine strategies to change eating behavior in IBD patients.

## Data Availability

Individual participant data will not be shared to preserve patient privacy.

## ACKNOWLEDGEMENTS

Icons for figures were created with Noun Project (creators: Amethyst Studio, Adrien Coquet, Muhammad Ikhsan, khaerul ilyas, Maria Kislitsina, Lihum Studio, popcomarts, Dio Sugiharto, and Stephen JB Thomas) under license CC BY 3.0.

## INFORMED CONSENT

This study was approved by the University of Miami Institutional Review Board (protocol number 20190548). All participants provided written informed consent before participating in this study.

## Abbreviations

ARFID (avoidant/restrictive food intake disorder), ASA24 (Automated Self-Administered 24-hour Dietary Assessment Tool), BMI (body mass index), CD (Crohn’s disease), CRP (C-reactive protein), HBI (Harvey-Bradshaw index), HEI (healthy eating index), HHC (healthy household control), HIPAA (Health Insurance Portability and Accountability Act), IBD (inflammatory bowel disease), JAK (Janus kinase), MAFLD (metabolic dysfunction-associated fatty liver disease), PRO2 (Patient-Reported Outcomes-2), PROMIS-29 (Patient-Reported Outcomes Measurement Information System-29), QoL (quality of life), SAA (serum amyloid A), sCDAI (Short Crohn’s Disease Activity Index), SFA (saturated fatty acid), SCFA (short-chain fatty acid), TLR4 (Toll-like receptor 4), TNFα (tumor necrosis factor alpha), UC (ulcerative colitis), VTE (venous thromboembolism)

## CRediT Authorship Contributions

Maria T. Abreu (Conceptualization: Lead; Funding acquisition: Lead; Methodology: Lead; Resources: Lead; Supervision: Lead; Validation: Lead; Writing – review & editing: Lead)

Maria A. Quintero (Conceptualization: Equal; Data curation: Supporting; Project administration: Lead; Acquired patient data and samples: Lead)

Luis Garces (Data curation: Supporting; Acquired patient data and samples: Lead) Hajar Hazime (Investigation: Equal)

Rose Killian (Formal analysis: Lead)

Katerina M. Faust (Visualization: Lead; Writing – original draft: Lead; Writing – review & editing: Contributing)

Payton Mendygral (Formal analysis: Supporting)

Judith Pignac-Kobinger (Conceptualization: Supporting)

Cristiana Mangarelli (Acquired patient data and sample: Supporting) Ana M. Santander (Investigation: Equal)

Irina Fernández (Investigation: Equal)

Norma Solis (Resources: Supporting; Acquired patient data and samples: Supporting) Mailenys Ortega (Resources: Supporting; Acquired patient data and samples: Supporting) Oriana M. Damas (Resources: Supporting; Acquired patient data and samples: Supporting)

Siobhan Proksell (Resources: Supporting; Acquired patient data and samples: Supporting) David H. Kerman (Resources: Supporting; Acquired patient data and samples: Supporting) Amar R. Deshpande (Resources: Supporting; Acquired patient data and samples: Supporting) Jennifer Seminerio (Resources: Supporting; Acquired patient data and samples: Supporting) Jana A.L. Hashash (Methodology: Supporting)

Philip Harvey (Methodology: Supporting) Ingrid Barrera (Methodology: Supporting) Tracy Crane (Methodology: Supporting)

## Data Transparency Statement

Individual participant data will not be shared to preserve patient privacy.

