## Supplementary Materials for "A high-fiber, low-fat diet improves the symptoms and metabolic profile of patients with Crohn’s disease"

Supplementary Methods

We recruited CD patients with mild to moderate symptoms (**Supplementary Table 1**). Patients taking biologics (anti-TNFs, anti-IL-12/23, vedolizumab) or immunomodulators (AZA, 6-MP, or methotrexate) had to have been on a stable dose for one month before screening. Patients taking oral mesalamines, sulfasalazine, or small molecules (S1P receptor agonists, JAK inhibitors), had to be on a stable dose for 2 weeks before screening. For inclusion, patients could receive no more than the equivalent of 20 mg of prednisone; they were allowed to taper their steroid dose as per their treating physician.

We excluded individuals with celiac disease or food allergies because these conditions would have complicated the catering process. We also excluded CD patients with an ileostomy or a colectomy because these conditions lack a validated symptom score. Pregnant and lactating women were excluded.

**Supplementary Table 1.** CD patient inclusion and exclusion criteria.

| **Criteria** | **Details** |
|---|---|
| *Inclusion criteria* | |
| Aged 18-70 years | |
| Documented diagnosis of Crohn's disease | sCDAI score < 400 |
| Living with someone with whom they share meals | |
| Available laboratory measurements within 3 months of screening | Hemoglobin, albumin creatine, BUN, eGFR, and a complete metabolic panel (CMP) |
| If on biologics or immunomodulators, stable dose for at least 1 month before screening | Anti-TNFs, anti-IL-12/23, vedolizumab, or immunosuppressants (AZA, 6-MP, or methotrexate) |
| Stable dose for at least 2 weeks before screening | Oral 5-aminosalicylates, mesalamine, or sulfasalazine |
| No antibiotic or probiotic use within 2 weeks prior to screening | |
| If taking steroids, use within the specified doses | No more than 20 mg of prednisone or 9 mg of budesonide per day<br><br>If clinically indicated, tapering of steriods was allowed after 4 weeks of the intervention; tapering had to involve a decrease of no more than 2.5 mg/week for prednisone and no more than 3 mg/week for budesonide |
| *Exclusion criteria* | |
| Gastrointestinal conditions or abnormalities | Ulcerative colitis, celiac disease, abdominal abscess, symptomatic intestinal stricture, or altered anatomy (prior total colectomy or proctocolectomy or anticipated colectomy during the study period and presence of ileal pouch or ostomy) |

| | |
|---|---|
| Clinically meaningful laboratory abnormalities | Unstable or uncontrolled medical disorder, colonic dysplasia or adenoma, malignant neoplasms, or toxic megacolon |
| Laboratory abnormalities within 3 months prior to screening or during the screening period | Hemoglobin < 8.0 g/dl or albumin < 2.8g/dl |
| Use of the following drugs within 2 months before screening | Cyclosporine, mycophenolate mofetil, sirolimus, thalidomide, or tacrolimus |
| Use of intravenous corticosteroids* within 2 weeks before screening, during screening, or during the study period | *Except as premedication for anti-TNFs |
| Daily use of steroids at the time of screening | Prednisone > 20 mg or budenoside > 9 mg |
| Use of total parenteral nutrition at screening or during the study period | |
| Uncooperative behavior or any condition that could make the patient potentially noncompliant to the study procedure | |
| Other significant or life-threatening comorbidities that could be negatively impacted by the low-fat/high-fiber diet | |
| Antibiotic use during the study period | |
| Pregnancy or lactation | |
| Known allergy to tree nuts or peanuts | |

**Group details**

Group 1 received a one-time 45-minute diet counseling session as well as a two-page diet guideline reference recommending a low-fat, high-fiber diet. Group 2 and Group 3 received the *Mi*-IBD diet through catered meals delivered to their homes for 8 weeks; catered meals were also provided to another individual without any gastrointestinal disease in the same household as patients in Group 3 (the HHC group).

**Questionnaires**

*Automated Self-Administered 24-hour Dietary Assessment Tool (ASA24)*[1]: This self-report tool is used to assess micronutrients, macronutrients, and total energy consumed based on participant recall of foods consumed over the past 24 hours.

*Healthy eating index*: This index is used to measure diet quality relative to the United States Department of Agriculture (USDA) recommendations in the *Dietary Guidelines for Americans*[2] and evaluates 13 dietary components, including total fruits, total vegetables, whole grains, refined grains, and fatty acids.

*Patient-Reported Outcomes-2 (PRO2):* This two-item self-report questionnaire is used to assess the severity of two symptoms of CD, stool frequency (the number of liquid or soft stools) and abdominal pain (from 0=*none* to 3=*severe*), over the past week[3]. Higher scores indicate greater severity of CD symptoms.

*Short Crohn's Disease Activity Index (sCDAI):* This questionnaire[4] assesses three symptoms of CD: the number of liquid or soft stools, severity of abdominal pain (from 0=*none* to 4=*severe*), and general well-being (from 0=*generally well* to 4=*terrible*) over the past seven days. The total scores on the short version[5] were designed to parallel those derived using the full CDAI. Specifically, sCDAI scores <150 indicate remission, while those ≥150, 221, and 451 indicate mild, moderate, and severe disease, respectively.

*Harvey-Bradshaw Index (HBI):* This index[6] contains five items: general well-being on the day before (from 0=*very well* to 4=*terrible*), abdominal pain on the day before (from 0=*none* to 3=*severe*), the number of liquid or soft stools per day, the presence of an abdominal mass (from 0=*none* to 3=*definite and tender*), and complications (1 point assigned per complication present. The total score ranges from 0 to 18, and the following thresholds are used to categorize CD severity: <5 (remission), 5-7 (mild disease), 8-16 (moderate disease), and >16 (severe disease).

**Catering Companies**

Four catering companies provided the catered food adhering to the *Mi*-IBD diet. The companies are as follows: The Healthy Chef, The Healthy Food Path, Lyfestyle Catering Inc., and Trifecta. Lyfestyle Catering Inc. provided catered food for the local area (Miami, FL). Trifecta provided catered food for participants within the state of Florida. The Healthy Chef and The Healthy Food Path provided catered food for participants in other states; the former closed during the COVID-19 pandemic, but the latter was started by the same chef and thus provided essentially the same catered food, differing only in ownership.

*Sample menu*:
Breakfast – egg whites with turkey bacon and Brussel sprouts
Lunch – crabcake with sumac carrots and chickpeas
Dinner – teriyaki salmon with green beans and quinoa

**Catered food: Intake data and preprocessing steps**

A research dietitian crafted all menus for the catered food, which were logged in the Nutrihand platform (http://www.nutrihand.com), a Health Insurance Portability and Accountability Act (HIPAA)-compliant electronic food diary. This platform facilitated recording food intake and monitoring adherence to the prescribed diet. The meals and snacks met 100% of the participants' nutritional needs, considering their BMI, age, gender, and physical activity levels. Calories were tailored to each participant's expenditures according to the Harris–Benedict equation to facilitate weight maintenance. The food was prepared at the catering facility and delivered to the participants' homes. Participants were trained to use Nutrihand to document all daily food consumption from their assigned menus and were instructed about foods that were appropriate substitutions for the catered meal to ensure the consumption of food with a similar nutritional profile.

We monitored the fidelity of the *Mi*-IBD diet in various ways. First, we tracked food delivery via receipts provided by the catering company. Second, the study dietician contacted participants on a weekly basis to document and resolve problems with the catered foods or catering procedures over the eight-week period. Third, participants were instructed to record all the food they ate in a web-based food diary (Nutrihand), which was pre-populated with the catered meal information. Fourth, adherence was assessed with the following question "Did you eat the food?". The response options were *always, usually, not usually,* and *seldom/never.* Additionally, participants were asked to rate the quality of the catered food on a scale from 1 (very poor) to 10 (excellent).

Participants also completed the Automated Self-Administered 24-hour Dietary Assessment Tool (ASA24) at baseline and week 8 (Group 1 only). At each time point, participants completed the ASA24 for two weekdays and one weekend day. Before the commencement of the diet intervention (baseline), participants received comprehensive instructions from a registered dietitian on utilizing the ASA24. Dietary intake data were downloaded from the ASA24 researcher workbench. For each daily observation of caloric intake, a participant-specific z-score was calculated using the caloric intake of all observations for that participant. Z-scores above or below 2 were excluded because these days were identified as possibly containing erroneous dietary entries. Next, a weekly average was calculated for each participant for each week in the study period. To account for week-to-week fluctuations in overall caloric intake, the macronutrient intake values were divided by the caloric intake for that week to create adjusted nutrient intake values.

Supplementary Results

**Dietary Adherence**

Almost all patients (96%) reported that they *always* or *usually* ate the catered food when asked; this adherence rate was similar across groups receiving catering (Group 2: 96% (26/27 participants), Group 3: 96% (23/24 participants), HHCs: 96% (23/24 participants). In general, participants enjoyed the catered food, with a mean rating of 7.57 (SD = 1.56) out of 10.

**Supplementary Table 1.** Dietary intake results. Omnibus tests were performed if analyses included three or more groups and consisted of one-way ANOVAs and Welch's ANOVAs, as appropriate. Pairwise comparisons were performed with Student's t test and are presented as unadjusted p values except for the intake of protein, total fat, saturated fat, and fiber (bold in these comparisons indicates a significant finding at an adjusted alpha threshold of 0.05/4 = 0.0125.

| **Variable** | **Omnibus test** | **Pairwise comparisons** |
|---|---|---|
| Protein | | |
| Between-group differences: Baseline | F(3, 50.8) = 1.34, p = 0.27 | -- |
| Within-group differences: Baseline vs. week 8 | -- | Group 1: t(22) = -1.41, p = 0.14 |
| | | Group 2: t(25) = -4.84, p = **$5.7*10^{-5}$** |
| | | Group 3: t(22) = -3.93, **p = $7.0*10^{-4}$** |
| | | HHCs: t(23) = -6.36, **p = $1.7*10^{-6}$** |
| Total fat | | |
| Between-group differences: Baseline | F(3, 92) = 0.22, p = 0.88 | -- |
| Within-group differences: Baseline vs. week 8 | -- | Group 1: t(22) = 0.82, p = 0.42 |
| | | Group 2: t(25) = 9.75, p = **$5.0*10^{-10}$** |
| | | Group 3: t(22) = 9.80, p = **$1.7*10^{-9}$** |
| | | HHCs: t(23) = 6.25, p = **$2.2*10^{-6}$** |
| Saturated fat | | |
| Between-group differences: Baseline | F(3, 92) = 1.27, p = 0.29 | -- |
| Within-group differences: Baseline vs. week 8 | -- | Group 1: t(22) = 1.43, p = 0.17 |
| | | Group 2: t(25) = 8.23, p = **$1.4*10^{-8}$** |
| | | Group 3: t(22) = 6.75, p = **$9.0*10^{-7}$** |
| | | HHCs: t(23) = 7.87, p = **$5.6*10^{-8}$** |
| Fiber | | |
| Between-group differences: Baseline | F(3, 92) = 0.11, p = 0.95 | -- |
| Within-group differences: Baseline vs. week 8 | -- | Group 1: t(22) = -1.66, p = 0.11 |
| | | Group 2: t(25) = -11.65, p = **$1.4*10^{-11}$** |
| | | Group 3: t(22) = -9.89, p = **$1.5*10^{-9}$** |
| | | HHCs: t(23) = -16.1, p = **$5.3*10^{-14}$** |
| Sugar | | |
| Between-group differences: Baseline | F(3, 92) = 0.36, p = 0.78 | |
| | -- | Group 1: t(22) = -0.45, p = 0.66 |
| | | Group 2: t(25) = -2.60, p = 0.02 |

| | | |
|---|---|---|
| Within-group differences: Baseline vs. week 8 | | Group 3: t(22) = -3.95, $p = 7.0*10^{-4}$ |
| | | HHCs: t(23) = -3.11, p = 0.005 |
| Omega 3:6 fatty acid ratio | | |
| Between-group differences: Baseline | F(3, 49.0) = 0.82, p = 0.5 | -- |
| Within-group differences: Baseline vs. week 8 | -- | Group 1: t(22) = 0.77, p = 0.45 |
| | | Group 2: t(25) = 7.50, $p = 1.0*10^{-7}$ |
| | | Group 3: t(22) = 5.00, $p = 5.2*10^{-5}$ |
| | | HHCs: t(22) = 9.91, $p = 9.0*10^{-10}$ |
| HEI total score | | |
| Between-group differences: Baseline | F(3, 93) = 0.43, p = 0.73 | -- |
| Within-group differences: Baseline vs. week 8 | -- | Group 1: t(22) = 2.45, p = 0.023 |
| | | Group 2: t(25) = -10.3, $p = 1.7*10^{-10}$ |
| | | Group 3: t(22) = -10.5, $p = 4.8*10^{-10}$ |
| | | HHCs: t(23) = -11.5, $4.8*10^{-10}$ |
| HEI total vegetables | | |
| Between-group differences: Baseline | F(3, 92) = 0.629, p = 0.598 | -- |
| Between-group differences: Week 8 | F(3, 48.5) = 13.09, p = **$2.2*10^{-6}$** | Group 1 vs. Group 2: $p = 1.9*10^{-10}$ |
| | | Group 1 vs. Group 3: $p = 5.1*10^{-10}$ |
| | | Group 1 vs. HHCs: $9.7*10^{-12}$ |
| Within-group differences: Baseline vs. week 8 | -- | Group 1: t(22) = -0.96, p = 0.35 |
| | | Group 2: $p = 5*10^{-6}$ |
| | | Group 3: $p = 9.6*10^{-4}$ |
| | | HHCs: $p = 1.1*10^{-4}$ |
| HEI greens & beans | | |
| Between-group differences: Baseline | F(3, 50.8) = 0.3, p = 0.826 | -- |
| Between-group differences: Week 8 | F(3, 48.5) = 13.09, p = **$2.2*10^{-6}$** | Group 1 vs. Group 2: $p = 1.7*10^{-7}$ |
| | | Group 1 vs. Group 3: $p = 8.7*10^{-7}$ |
| | | Group 1 vs. HHCs: $p = 2.2*10^{-7}$ |
| Within-group differences: Baseline vs. week 8 | -- | Group 1: t(22) = 0.85, p = 0.41 |
| | | Group 2: $p = 1.7*10^{-6}$ |
| | | Group 3: $p = 1.4*10^{-6}$ |
| | | HHCs: $p = 1.2*10^{-5}$ |
| HEI total fruit | | |
| Between-group differences: Baseline | F(3, 92) = 1.60, p = 0.195 | -- |
| Between-group differences: Week 8 | F(3, 48.5) = 13.09, p = **$2.2*10^{-6}$** | Group 1 vs. Group 2: $p = 3.6*10^{-5}$ |
| | | Group 1 vs. Group 3: $p = 2.2*10^{-6}$ |
| | | Group 1 vs. HHCs: $p = 1.2*10^{-5}$ |
| | -- | Group 1: t(22) = -0.58, p = 0.57 |

| | | |
|---|---|---|
| Within-group differences: Baseline vs. week 8 | | Group 2: $p = 1.4*10^{-6}$ |
| | | Group 3: $p = 2*10^{-7}$ |
| | | HHCs: $p = 5.3*10^{-5}$ |
| HEI whole fruit | | |
| Between-group differences: Baseline | $F(3, 92) = 1.18$, $p = 0.323$ | -- |
| Between-group differences: Week 8 | $F(3, 48.5) = 13.09$, $p =$ **$2.2*10^{-6}$** | Group 1 vs. Group 2: $p = 4.8*10^{-6}$ |
| | | Group 1 vs. Group 3: $p = 2.1*10^{-7}$ |
| | | Group 1 vs. HHCs: $p = 5.6*10^{-6}$ |
| Within-group differences: Baseline vs. week 8 | -- | Group 1: $t(22) = -0.61$, $p = 0.55$ |
| | | Group 2: $p = 4*10^{-7}$ |
| | | Group 3: $p = 4*10^{-7}$ |
| | | HHCs: $p = 1.1*10^{-5}$ |
| HEI whole grain | | |
| Between-group differences: Baseline | $F(3, 92) = 0.108$, $p = 0.955$ | -- |
| Between-group differences: Week 8 | $F(3, 48.5) = 13.09$, $p =$ **$2.2*10^{-6}$** | Group 1 vs. Group 2: $p = 5.1*10^{-10}$ |
| | | Group 1 vs. Group 3: $p = 4.9*10^{-10}$ |
| | | Group 1 vs. HHCs: $p = 1.2*10^{-9}$ |
| Within-group differences: Baseline vs. week 8 | -- | Group 1: $t(22) = 2.24$, $p = 0.04$ |
| | | Group 2: $p = 4.5*10^{-8}$ |
| | | Group 3: $p = 5*10^{-7}$ |
| | | HHCs: $p = 3*10^{-11}$ |
| HEI refined grain | | |
| Between-group differences: Baseline | $F(3, 92) = 0.725$, $p = 0.54$ | -- |
| Between-group differences: Week 8 | $F(3, 48.5) = 13.09$, $p =$ **$2.2*10^{-6}$** | Group 1 vs. Group 2: $p = 1.4*10^{-12}$ |
| | | Group 1 vs. Group 3: $p = 5.2*10^{-12}$ |
| | | Group 1 vs. HHCs: $p = 2.6*10^{-11}$ |
| Within-group differences: Baseline vs. week 8 | -- | Group 1: $t(22) = 1.63$, $p = 0.12$ |
| | | Group 2: $p = 1*10^{-5}$ |
| | | Group 3: $3.7*10^{-6}$ |
| | | HHCs: $p = 5.3*10^{-5}$ |
| HEI seafood & plant protein | | |
| Between-group differences: Baseline | $F(3, 92) = 0.248$, $p = 0.862$ | -- |
| Between-group differences: Week 8 | $(F(3, 48.5) = 13.09$, $p =$ **$2.2*10^{-6}$** | Group 1 vs. Group 2: $p = 5.3*10^{-4}$ |
| | | Group 1 vs. HHCs: $p = 0.006$ |
| Within-group differences: Baseline vs. week 8 | -- | Group 1: $t(22) = 2.71$, $p = 0.013$ |
| | | Group 2: $p = 3.3*10^{-5}$ |
| | | Group 3: $p = 7.3*10^{-5}$ |
| | | HHCs: $p = 1.1*10^{-4}$ |

Abbreviations: HEI = healthy eating index, HHC = healthy household control.

**Supplementary Table 2.** CD symptom severity. Omnibus tests were performed if analyses included three or more groups and consisted of one-way ANOVAs and Welch's ANOVAs, as appropriate. Pairwise comparisons were performed with Student's t test and are presented with unadjusted p values unless otherwise specified. Bold indicates significant p values.

| **Variable** | **Omnibus test** | **Pairwise comparisons** |
|---|---|---|
| HBI | | |
| Between-group differences: Baseline | F(2, 69) = 0.18, p = 0.83 | -- |
| Between-group differences: Week 8 | F(2, 69) = 0.02, p = 0.98 | -- |
| Within-group differences: Baseline vs. week 8 | -- | Group 1: t(22) = 1.25, p = 0.22 |
| | | Group 2: t(25) = 1.65, p = 0.11 |
| | | Group 3: t(22) = 0.96, p = 0.35 |
| PRO2 | | |
| Between-group differences: Baseline | F(2, 69) = 0.24, p = 0.79 | -- |
| Between-group differences: Week 8 | F(2, 43.3) = 0.59, p = 0.56 | -- |
| Within-group differences: Baseline vs. week 8 | -- | Group 1: t(22) = 1.46, p = 0.16 |
| | | Group 2: t(25) = 0.49, p = 0.63 |
| | | Group 3: t(22) = 2.10, p = **0.05** |
| sCDAI | | |
| Between-group differences: Baseline | F(2, 69) = 0.05, p = 0.95 | -- |
| Between-group differences: Week 8 | F(2, 43.3) = 0.63, p = 0.54 | -- |
| Within-group differences: Baseline vs. week 8 | -- | Group 1: t(22) = 2.02, p = 0.06 |
| | | Group 2: t(25) = 1.21, p = 0.24 |
| | | Group 3: t(22) = 2.51, p = **0.02** |

Abbreviations: HBI = Harvey-Bradshaw index, PRO2 = 2-item Patient-Reported Outcomes, sCDAI = Short Crohn's Disease Activity Index.

**Supplementary Table 3.** Inflammatory biomarker levels. Omnibus tests were performed if analyses included three or more groups and consisted of one-way ANOVAs and Welch's ANOVAs, as appropriate. Pairwise comparisons were performed with Student's t test and are presented with unadjusted p values unless otherwise specified. Bold indicates significant p values.

| Variable | Omnibus test | Pairwise comparisons |
|---|---|---|
| CRP | | |
| Between-group differences: Baseline | F(3, 43.13) = 1.59, p=0.205 | -- |
| Between-group differences: Week 8 | F(3, 49.11) = 0.39, p=0.764 | -- |
| Within-group differences: Baseline vs. week 8 | -- | Group 1: t(19)=1.22, p=0.24 |
| | | Group 2: t(25)=-1.16, p=0.26 |
| | | Group 3: t(20)=0.94, p=0.36 |
| | | HHCs: t(22)=1.20, p=0.24 |
| FCP | | |
| Between-group differences: Baseline | F(3, 45.00) = 1.67, p=0.186 | -- |
| Between-group differences: Week 8 | F(3, 49.11) = 6.42, p=**0.001** | No significant pairwise comparisons |
| Within-group differences: Baseline vs. week 8 | -- | Group 1: t(18)=0.663, p=0.52 |
| | | Group 2: t(24)=-1.87, p=0.07 |
| | | Group 3: t(20)=0.43, p=0.68 |
| | | HHCs: t(19)=1.52, p=0.15 |
| SAA | | |
| Between-group differences: Baseline | F(3, 49.77) = 0.28, p=0.842 | -- |
| Between-group differences: Week 8 | F(3,47.36) = 0.93, p=0.433 | -- |
| Within-group differences: Baseline vs. week 8 | -- | Group 1: t(22)=0.05, p=0.96 |
| | | Group 2: t(21)=-0.15, p=0.88 |
| | | Group 3: t(20)=-1.02, p=0.32 |
| | | HHCs: t(20)=1.73, p=0.10 |

Abbreviations: CRP = C-reactive protein, FCP = fecal calprotectin, HHC = healthy household control, SAA = serum amyloid A.

### SUPPLEMENTARY FIGURES

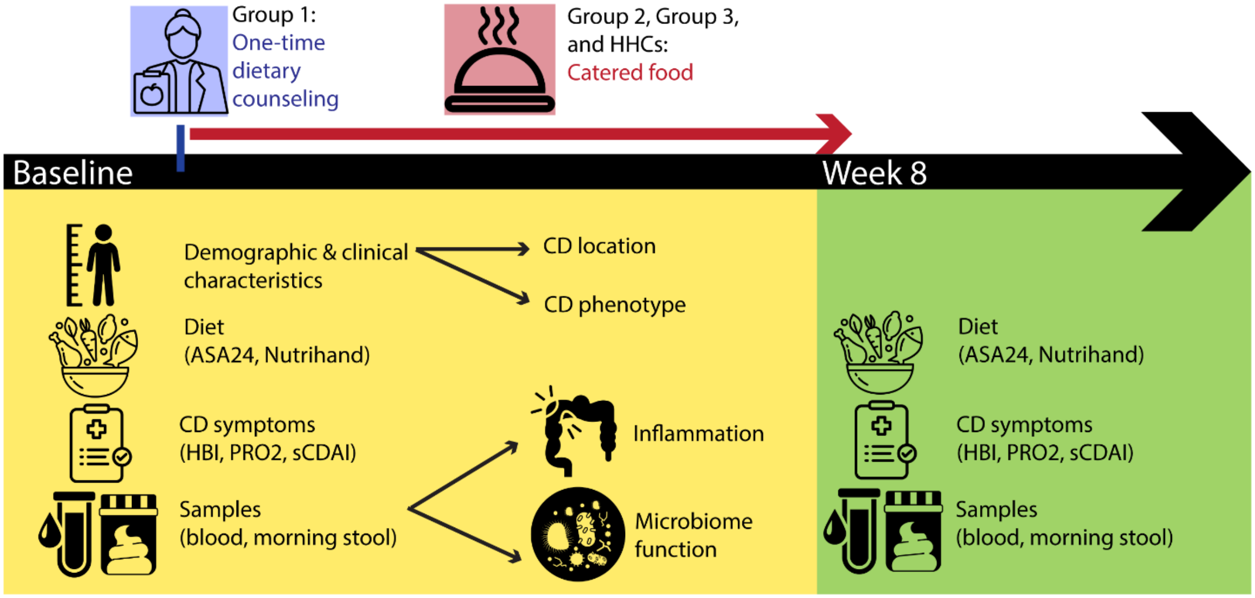


**Supplementary Figure 1.** Schematic of data collected at baseline vs. week 8.

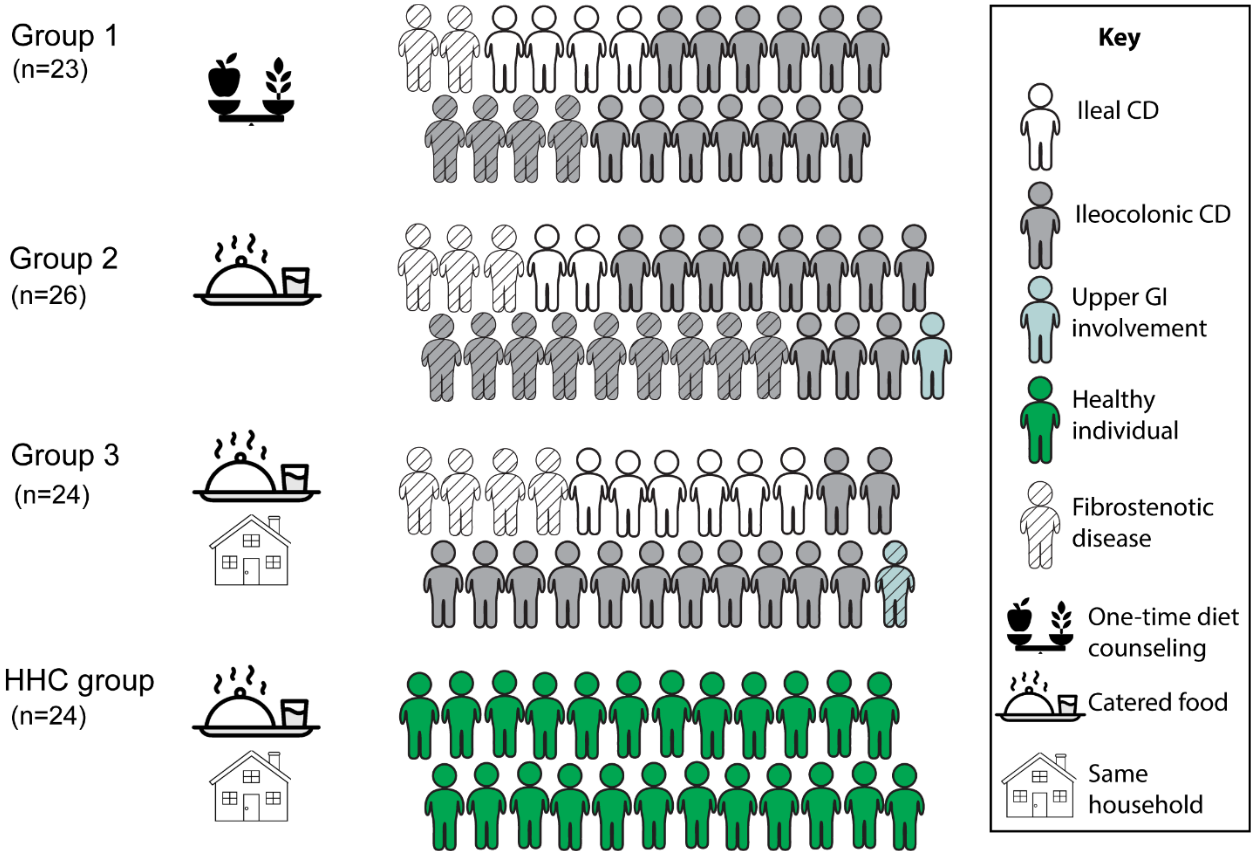


**Supplementary Figure 2.** Group composition. Abbreviations: CD = Crohn's disease, GI = gastrointestinal, HHC = healthy household control.

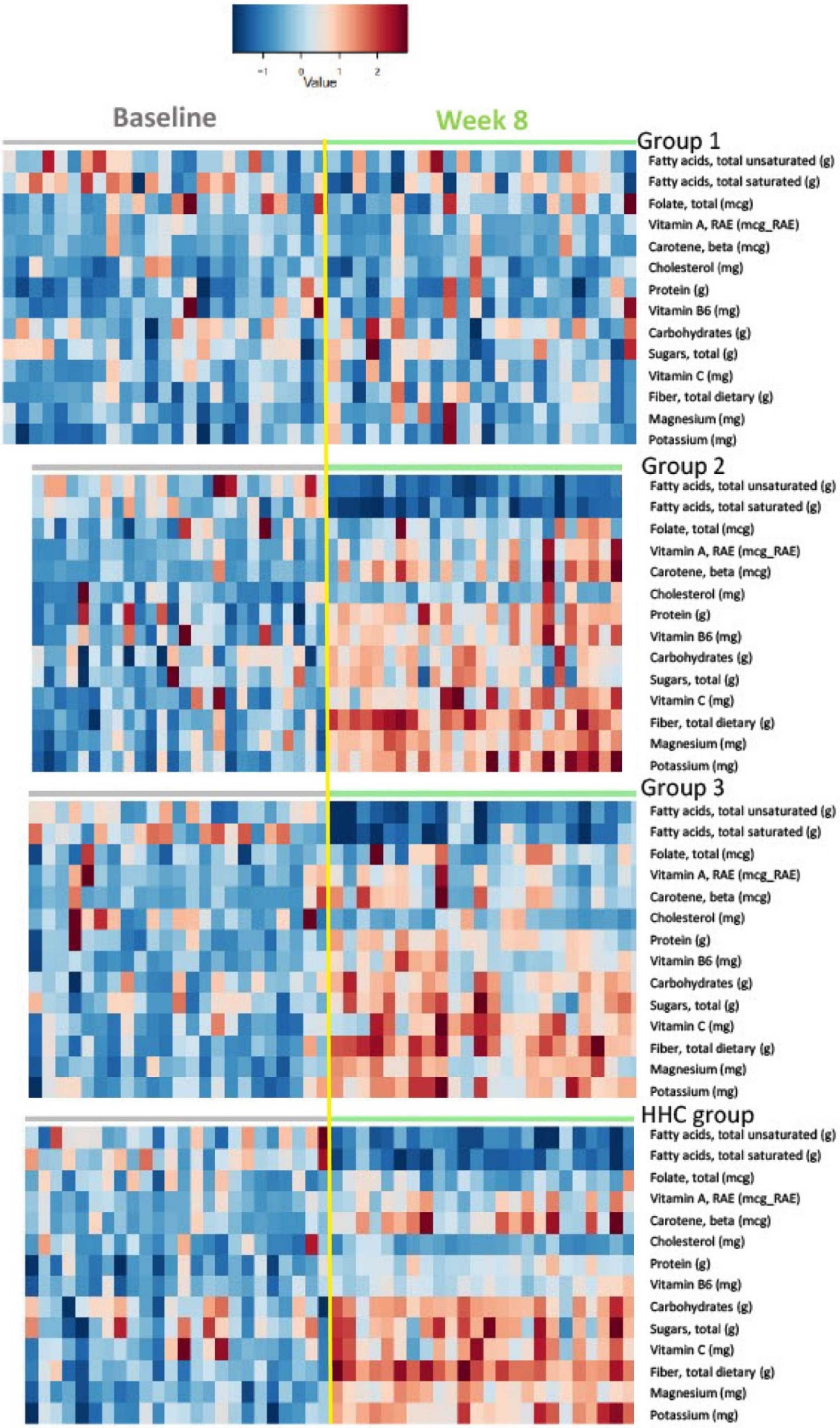

-1
0
1
2
Value
Baseline
Week 8
Group 1
Fatty acids, total unsaturated (g)
Fatty acids, total saturated (g)
Folate, total (mcg)
Vitamin A, RAE (mcg_RAE)
Carotene, beta (mcg)
Cholesterol (mg)
Protein (g)
Vitamin B6 (mg)
Carbohydrates (g)
Sugars, total (g)
Vitamin C (mg)
Fiber, total dietary (g)
Magnesium (mg)
Potassium (mg)
Group 2
Fatty acids, total unsaturated (g)
Fatty acids, total saturated (g)
Folate, total (mcg)
Vitamin A, RAE (mcg_RAE)
Carotene, beta (mcg)
Cholesterol (mg)
Protein (g)
Vitamin B6 (mg)
Carbohydrates (g)
Sugars, total (g)
Vitamin C (mg)
Fiber, total dietary (g)
Magnesium (mg)
Potassium (mg)
Group 3
Fatty acids, total unsaturated (g)
Fatty acids, total saturated (g)
Folate, total (mcg)
Vitamin A, RAE (mcg_RAE)
Carotene, beta (mcg)
Cholesterol (mg)
Protein (g)
Vitamin B6 (mg)
Carbohydrates (g)
Sugars, total (g)
Vitamin C (mg)
Fiber, total dietary (g)
Magnesium (mg)
Potassium (mg)
HHC group
Fatty acids, total unsaturated (g)
Fatty acids, total saturated (g)
Folate, total (mcg)
Vitamin A, RAE (mcg_RAE)
Carotene, beta (mcg)
Cholesterol (mg)
Protein (g)
Vitamin B6 (mg)
Carbohydrates (g)
Sugars, total (g)
Vitamin C (mg)
Fiber, total dietary (g)
Magnesium (mg)
Potassium (mg)

**Supplementary Figure 3.** Heatmaps of nutrient intake from baseline to week 8 based on ASA24 data. Colors reflect intake at various time points: cool colors = low intake of a specific nutrient, warm colors = high intake of a specific nutrient. Rows represent nutrients, columns represent individual participants.

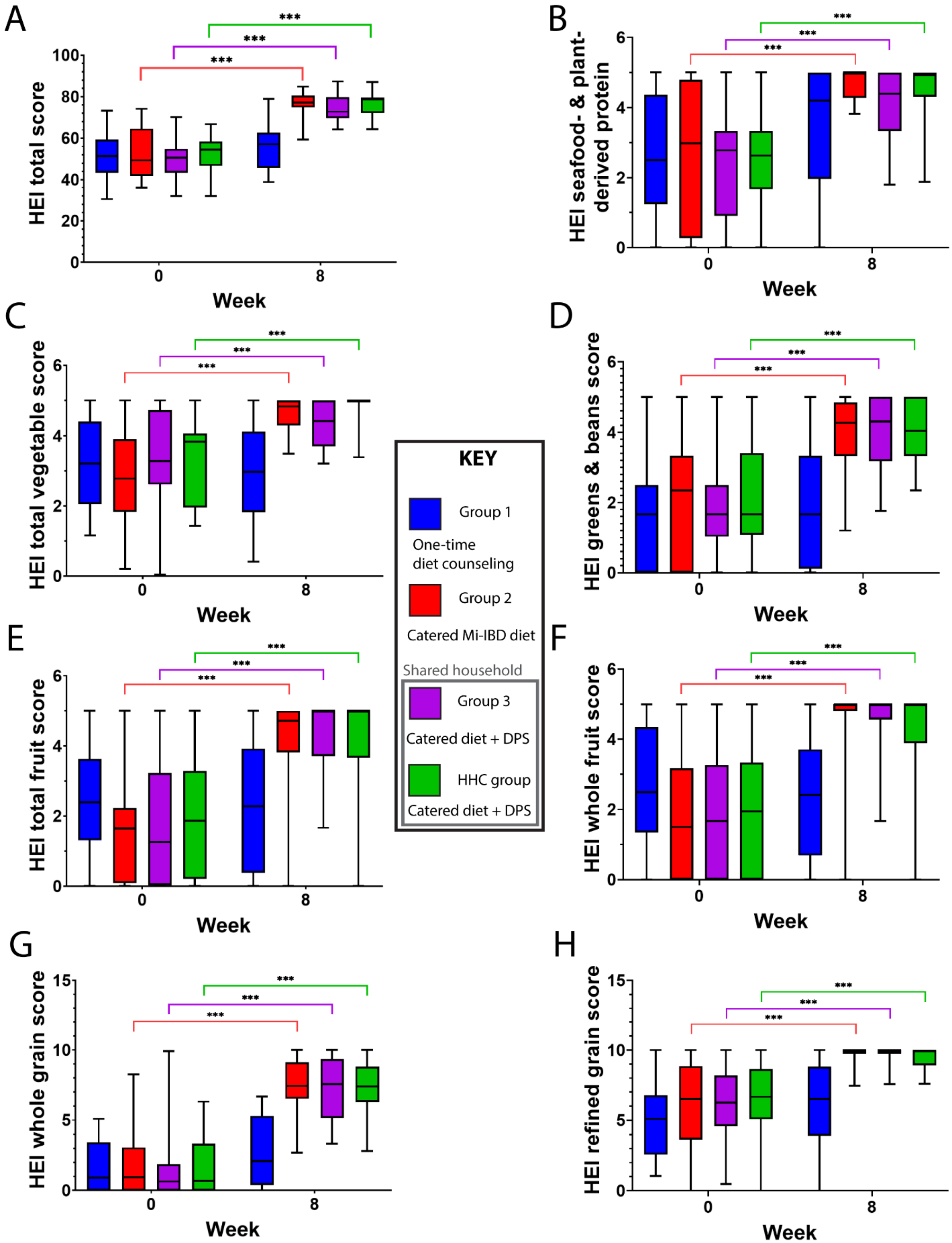


**Supplementary Figure 5.** Participants who received the catered food (Group 2, Group 3, and the HHC group) exhibited improvements in dietary quality. A) HEI total score. B) HEI seafood-

and plant-derived protein score. C) HEI total vegetable score. D) HEI greens and beans score. E) HEI total fruit score. F) HEI whole fruit score (i.e., all fruit excluding fruit juice). G) HEI whole grain score. H) HEI refined grain score. Higher HEI scores in any category reflect better dietary. Abbreviations: HEI = healthy eating index (higher scores reflect a healthier diet for all components), HHC = healthy household control. Data were compared within groups with paired t-tests, and Bonferroni correction was applied. * $p < 0.05$, ** $p < 0.01$, *** $p < 0.001$.
